# Factors influencing COVID-19 vaccination decision-making among African American and Hispanic pregnant and postpartum women in Deep South

**DOI:** 10.1101/2023.07.20.23292951

**Authors:** Ran Zhang, Tiffany Byrd, Shan Qiao, Myriam E. Torres, Xiaoming Li, Jihong Liu

## Abstract

**Background:** COVID-19 vaccination is vital for ending the pandemic but concerns about its safety among pregnant and postpartum women, especially among African American (AA) and Hispanic women, persist. This study aims to explore factors that influence vaccination decision-making among AA and Hispanic pregnant and postpartum women through women’s experiences and maternal care providers’ (MCPs) observations.

**Methods:** From January and August 2022, we conducted semi-structured interviews with AA and Hispanic women and MCPs. Participants were recruited from obstetric and pediatric clinics in South Carolina, and all births took place after March 2020. Thematic analysis was employed for data analysis.

**Results:** The study involved 19 AA and 20 Hispanic women, along with 9 MCPs, and revealed both barriers and facilitators to COVID-19 vaccination. The factors that influence pregnant and postpartum women’s decision about COVID-19 vaccine uptake included: 1) awareness of health threats associated with COVID-19 vaccines, 2) vaccine availability and accessibility, 3) vaccine-related knowledge and exposure to misinformation, 4) concerns regarding pre-existing health conditions and potential side effects of COVID-19 vaccines, 5) emotional factors associated with vaccination decision-making processes, 6) concerns about the well-being of infants, 7) cultural perspectives, and 8) encouragement by trusted supporters.

**Conclusion:** Findings suggest that reliable information, social support, and trusted doctors’ advice can motivate COVID-19 vaccination. However, barriers such as misinformation, mistrust in the health care system, and fears related to potential side effects impede vaccination uptake among AA and Hispanic pregnant and postpartum women. Future interventions should target these barriers, along with health disparities, involve trusted doctors in outreach, and initiate vaccine conversations to promote vaccination among this population.

## 1. Introduction

The COVID-19 pandemic has caused a significant number of cases and deaths worldwide, making vaccination an important strategy to prevent severe adverse health outcomes. Although various health organizations, including the Centers for Disease Control and Prevention (CDC), the American College of Obstetricians and Gynecologists (ACOG), and the Society for Maternal-Fetal Medicine (SMFM), recommend COVID-19 vaccination for pregnant women, data indicate that vaccination coverage for pregnant women remains low compared to the general population (1, 2). Data from the National Immunization Survey (COVID-19 supplemental module) showed that among women aged 18-49 years, COVID-19 vaccination coverage was lower among pregnant women (45%) and breastfeeding women (51%) than non-pregnant women (65%) (3). This is concerning, as pregnant women have a higher risk of severe illness and mortality from COVID-19 and are at risk for adverse pregnancy outcomes, such as preterm delivery and stillbirth, compared with non-pregnant women (4, 5). These disparities highlight the need to investigate the factors influencing COVID-19 vaccination decision-making among pregnant women.

The decision-making process for COVID-19 vaccination is influenced by various factors, including limited available safety data on COVID-19 vaccine during pregnancy, lack of confidence in the safety and efficacy of the vaccine among health care providers and pregnant women, issues related to vaccine prioritization, access, and availability, as well as cultural and language barriers (3, 6, 7). Moreover, pregnant women are particularly concerned about the potential safety issues for their children associated with COVID-19 vaccination, which stems from the limited data on the effects of the vaccine on fetal development (8). These unresolved concerns about the safety and efficacy of the COVID-19 vaccine can lead to hesitancy in their decision-making process. The research on COVID-19 vaccination during pregnancy has significantly expanded (9, 10). However, the presence of inconsistent information from social media and health authorities during the launch and promotion phases of the vaccine, coupled with conflicting advice from medical and public health professionals, has led to confusion and the perpetuation of misinformation (11, 12).

The impact of these factors is particularly pronounced among African American (AA) and Hispanic pregnant women, who face historical and contemporary racism and discrimination within the health care system (13, 14). Such experiences have led to mistrust and suspicion among these groups, creating barriers to seeking health care and following medical advice (15). In addition, AA and Hispanic people are more likely to experience socioeconomic and structural barriers to health care, such as inadequate access to health care facilities and insurance coverage (16). Thus, in such a sociocultural context, the decision to receive COVID-19 vaccines during pregnancy becomes even more complex AA and Hispanic pregnant women, necessitating a careful consideration of the potential costs and benefits. There is a dearth of empirical studies focusing on factors affecting COVID-19 vaccination decision-making among AA and Hispanic pregnant and postpartum women. This study aims to explore the factors that influence vaccination decision-making among AA and Hispanic pregnant and postpartum women through women’s experiences and maternal care providers’ (MCPs) observations. By uncovering these factors, the study seeks to inform targeted interventions that promote vaccination and help reduce health disparities within these underserved populations.

## 2. Methods

### 2.1 Study Design and Data Collection

Between January and August 2022, we conducted a qualitative study as part of a larger project that examined maternal care utilization and psychosocial well-being among AA and Hispanic pregnant and postpartum women during the COVID-19 pandemic. Purposive sampling was employed to recruit postpartum women and MCPs. Participants were recruited through local obstetrics and gynecology clinics and community health organizations that primarily served low-income AA and Hispanic women in South Carolina. The eligibility criteria included residency in South Carolina, age of at least 18 years, self-identification as AA or Hispanic, and all births occurring after March 2020. In total, we conducted 48 interviews, including 19 AA women, 20 Hispanic women, and 9 MCPs. The semi-structured interviews were conducted via Zoom, by phone, or in-person by two researchers. We obtained informed consent from each participant, emphasizing their voluntary participation and right to withdraw at any time. Each interview lasted approximately 50 minutes and was audio recorded with each participant’s consent. Each participant received a $50 gift card upon completion of the interview. The study received ethical approval from the University of South Carolina Institutional Review Board (Pro00115169).

### 2.2 Data Analysis

The interviews were transcribed verbatim, and for interviews conducted in Spanish, one of the researchers translated them into English for analysis. Anonymized transcriptions were then reviewed and subjected to inductive thematic analysis by two researchers (17). Each researcher individually assigned descriptive codes to the data, identifying frequently used phrases and examining the context in which they were used (17). Any coding discrepancies were resolved through discussion and by re-reviewing the transcripts. In the process of analyzing the interview transcripts, we found that a notable topic mentioned by the participants was COVID-19 vaccination decision-making, which encompassed awareness of health threats associated with COVID-19 vaccines, vaccine availability and accessibility, vaccine-related knowledge and misinformation, and concerns regarding the well-being of both women and their infants.

### 2.3 Demographic Characteristics

All the postpartum women participated in the study gave birth after March 2020 at a hospital in South Carolina. At the time of interview, AA women were between 21 to 42 years old, had at least a high school diploma or general education diploma, 74% were not first-time mothers, and had experience with maternal care services before and during COVID-19. In terms of insurance coverage, 58% of AA women had their births paid by Medicaid, 37% paid by private insurance, and 5% paid by themselves. AA women resided mainly in Richland (32%) and Spartanburg Counties (26%). Hispanic women were between 19 to 41 years old, 50% did not have a high school diploma, all were immigrants to the US, and 75% were not first-time mothers. Regarding insurance coverage, 70% of Hispanic women had their births covered by Emergency Medicaid, 5% paid by Medicaid, 5% paid by a special program, and 20% paid by themselves. Hispanic women predominantly live in Richland (45%) and Lexington Counties (45%). The MCPs who were interviewed had a variety of occupations, including obstetric gynecologists, maternal fetal medicine specialists, nurses, and community doulas, consisting of eight females and one male. Five of the MCPs worked in clinics that primarily served AA patients (50-80%), and three MCPs worked in clinics that served more evenly mixed patient populations (both AA and Hispanic patients comprised 20-40%) (Table 1).

**Table 1.** Sociodemographic characteristics of the participants.

| *Characteristic* | *African American*<br>*Participant (%)*<br>*N=19* | *Hispanic*<br>*Participant (%)*<br>*N=20* |
| --- | --- | --- |
| **Age Range** |  |  |
| 18-25 | 3 (16%) | 5 (25%) |
| 26-35 | 12 (63%) | 8 (40%) |
| 36-50 | 4 (21%) | 7 (35%) |
| **Education** |  |  |
| Less than high school diploma |  | 10 (50%) |
| High school diploma or general<br>education development (GED) | 9 (47%) | 6 (30%) |
| Associate degree | 1 (5%) |  |
| Bachelor degree | 5 (26%) | 3 (15%) |
| Master degree | 4 (21%) |  |
| No education |  | 1 (5%) |
| **Marital Status** |  |  |
| Single | 2 (11%) | 6 (30%) |
| Married | 9 (47%) | 6 (30%) |
| Living with unmarried partner | 8 (42%) | 8 (40%) |
| **Number of Children** |  |  |
| 1 | 5 (26%) | 5 (25%) |
| 2 | 8 (42%) | 3 (15%) |
| 3 | 4 (21%) | 6 (30%) |
| 4 | 2 (11%) | 3 (15%) |
| >4 |  | 3 (15%) |
| **Insurance Coverage** |  |  |
| Medicaid | 11 (58%) | 1 (5%) |
| Private insurance | 7 (37%) |  |
| Self-pay | 1 (5%) | 4 (20%) |
| Emergency Medicaid |  | 14 (70%) |
| Other/program |  | 1 (5%) |
| **Place of Birth** |  |  |
| USA | 19 (100%) |  |
| Colombia |  | 4 (20%) |
| Mexico |  | 4 (20%) |
| Guatemala |  | 6 (30%) |
| Honduras |  | 4 (20%) |
| Costa Rica |  | 1 (5%) |
| Unknown |  | 1 (5%) |
| **County of Residence** |  |  |
| Darlington | 1 (5%) |  |
| Spartanburg | 5 (26%) |  |
| Colleton | 3 (16%) |  |
| Beaufort | 1 (5%) |  |
| Richland | 6 (32%) | 9 (45%) |
| Sumter | 2 (11%) |  |
| Lexington | 1 (5%) | 9 (45%) |
| Berkeley |  | 1 (5%) |
| Unknown |  | 1 (5%) |

| *Characteristic* | *Maternal Care*<br>*Provider*<br>*N=9* |
| --- | --- |
| **Age** |  |
| 26-35 | 1 |
| 36-45 | 3 |
| 46-55 | 3 |
| 56-65 | 2 |
| **Tenure at Current Practice/Hospital** |  |
| <1 year | 2 |
| 1-5 years | 4 |
| 6-10 years | 3 |
| **Job Title/Specialty Area** |  |
| Certified Midwife | 2 |
| Obstetrics and<br>Gynecology/Resident/Maternal<br>Fetal Medicine Specialist | 4 |
| Registered Nurse/Nurse<br>Practitioner | 3 |

Note: Percentages do not add up to 100% due to rounding.

## 3. Results

Based on AA and Hispanic women’s experiences and MCPs’ observations, the current study revealed several factors that influenced the women’s decision-making regarding COVID-19 vaccination with the following key themes emerged: 1) awareness of health threats associated with COVID-19, 2) vaccine availability and accessibility, 3) vaccine-related knowledge and exposure to misinformation, 4) concerns regarding pre-existing health conditions and potential side effects of COVID-19 vaccines, 5) emotional factors associated with vaccination decision-making processes, 6) concerns about the well-being of infants, 7) cultural perspectives, and 8) encouragement by trusted supporters.

### 3.1 Awareness of health threats associated with COVID-19

Participants in both the AA and Hispanic groups expressed a strong awareness of the health threats posed by COVID-19 to themselves and others. They displayed stress and fear regarding their vulnerability to contracting the virus.

> “… *during my pregnancy, the biggest concern for me, I was always nervous about catching COVID, because for some reason, last year, all you saw on the news was all these pregnant women going into a coma when they had COVID. So that stressed me out.*” (8)

> “… *worried about getting sick, there were a lot of you heard on the news about people getting COVID and being pregnant and either the mom died or the baby died, or the mom had the baby but the mom still didn’t make it. So it was very scary.*” (8)

> “*Too scared about getting COVID and we didn’t know how it was going to be. Fear of seeing other people but at the same time I didn’t want to see anyone for that fear.*” (9)

> “*I was afraid to get sick especially at the beginning of the pandemic.*” (5)

The news coverage of pregnant women experiencing severe outcomes from COVID-19 contributed to their anxiety. Participants were particularly concerned about the potential risks to their own health and the health of their infants. They expressed fear of getting sick and the possibility of adverse outcomes for both themselves and their babies. The participants also acknowledged the risk of COVID-19 transmission within their households, especially to school-aged children. The optional mask-wearing policy in schools added to their worries.

> “*Then my daughter’s in school. So I worry about that, you know, they don’t mandate masks wearing masks is optional in her school, she does wear masks, but it’s just stressful because I don’t want to get her sick, I don’t want her to get sick. And I definitely don’t want my little baby getting sick.*” (1)

> “*Yes, I was nervous about schools. Um just because, like I said, in m environment, I can control, but you started going to school, and then, wondering if my son had his mask on at the right times, did other people expose him to anything.*” (5)

> “*Yes, because my daughter was in school and was worried that she would infect me with COVID. Also going to prenatal appointments and get infected there and not knowing what happened.*” (6)

Participants expressed concern about their ability to control the exposure of their children to the virus and the potential consequences of attending prenatal appointments. They highlighted their efforts to maintain a safe home environment and limit exposure to individuals entering and leaving their households. The anxiety and nervousness experienced by the participants led to heightened precautions such as frequent handwashing and avoidance of crowded places.

> “*It was very nerve wrecking. Like, I was nervous about lifting up my shirt. Or as soon as I would come home from my doctor’s appointment, I’m not even gonna lie to you, I jumped right into that shower. And I scrubbed from head to toe. And so it was just, I think I was more paranoid more over OCD [obsessive-compulsive disorder] as well. With this pandemic, especially being pregnant and even now, when I go out, when I come back, I wash ‘cause I don’t want to get sick.*” (3)

> “*Yes, I am very anxious and nervous about her being exposed. She’s not in daycare ‘cause of that reason. Actually, my mother and my husband’s mother is the only people that watch her while we’re at work. So we try to limit her exposure.*” (6)

> “*With the pandemic going on, I was like real skeptical about going anywhere. ‘cause I didn’t want catch COVID with me being pregnant. So I was really anxious due to the fact you know, I got help from my family. But for the most part, I had to do mostly everything myself. So it was like, do I want to leave the house? Do I have to leave the house? I was really nervous because of COVID.*” (2)

> “*Yes, I was scared to go to the store because I was pregnant.*” (8)

> “*I was scared of everyone who got close to the baby because his defenses are low.*” (6)

### 3.2 Vaccine availability and accessibility

Both MCPs and women expressed their observations and experiences regarding the waiting period for vaccine development and distribution. Their responses reflect a sense of anticipation and the understanding that the availability of vaccines was crucial for providing adequate care and managing the pandemic. Their responses suggest a mixed sentiment, with some expressing anxiety and depression related to the vaccine’s availability, while others acknowledge the challenges faced in obtaining necessary supplies and appointments due to the impact of COVID-19.

> “… *early on, when the vaccines weren’t available everybody wanted the vaccine now, then, when the vaccines became available, more people didn’t want the vaccine. It was a two-edged sword, but it was another source of anxiety and depression.*” (7)

> “*But it was kind of like, we were just sort of waiting on a vaccine, to be able to provide the same level of care. And we talked about this in the clinic, too, it’s like, doctors are supposed to see sick patients, it was just that everybody was so scared, ‘cause we hadn’t seen something like this before. And young people were dying.*” (8)

> “*So yeah, I feel like COVID pushed, help pushed a lot of stuff back. And then they weren’t able to get some of the supplies they needed because of COVID and everything was back ordered. ‘Cause I know that happened to me a few times. With the vaccines, I couldn’t get my shots on certain days. Because everything was backed up due to COVID.*” (3)

MCPs also acknowledged the difficulties patients faced in accessing the COVID-19 vaccine at the point of service. They expressed a sense of frustration regarding the limited on-site accessibility of vaccines, which potentially hindered their ability to provide comprehensive care.

> “*The vaccines are not accessible on site.*” (4)

> “*They stopped doing the vaccines in our building, ‘cause the demand was not as high … now more people were getting sick and then the surge for vaccines increased. But I mean, we were sending our OB [obstetrics] patients… to go to a different building.*” (MCP 4)

### 3.3 Vaccine-related knowledge and exposure to misinformation

Some participants were hesitant to get vaccinated because of misinformation they heard or read. While this was a persistent challenge for MCPs trying to educate patients about the benefits and safety of vaccination, misinformation still led some patients to choose not to vaccinate themselves or their children.

> “… *trying to educate them in terms of the vaccines, I mean, that’s just been a constant battle. You know, trying to help them understand that it’s safe, and it will protect you and the baby. But all the other misinformation that’s out there, and that’s definitely been a challenge and a battle, I will say, a lot more of our patients have gotten vaccinated than I really expected to see.*” (2)

> “… *when new things come, … when the vaccine did come out, then a lot of people did a lot of reading, my husband is like ‘let me find out about this first’. And I am too … I definitely wanted to know … first if anybody else any other pregnant person had taken the vaccine, and you read a couple things that are good. And then you come across that one thing. Mom doesn’t make it baby dies from vaccine and we were like, absolutely not.*” (8)

Skepticism played a significant role in the decision-making process of participants who chose not to vaccinate themselves. Both AA participants and MCPs highlighted skepticism towards the vaccine, the rapid development process, and concerns about government involvement.

> “*I’ve never been the type to take the vaccines anyway. But like looking into it, they just made that too fast I feel like, so I’m not saying I’m not, but I’m not doing that now. Like, I’ll just watch how everybody else … I’ll just keep taking my vitamins, be cleaning and all that stuff, staying out the way.*” (7)

> “*It actually did, because it was such the unknown and at the point of my pregnancy, it was just approved for pregnant women. … Is it safe for me to get it? Is it not okay? And I think I was about seven months pregnant, so I was so far long that I had no complications. Do I want to risk taking the vaccine and create a complication and I’ve been okay this whole time? So a lot of stress with that. Just going back and forth with the doctor’s asking questions on how it would affect the baby. How would it affect me in my pregnancy? Just a lot of going back and forth a lot of stress and anxiety about just that alone.*” (5)

> “… *as the pandemic progressed, there was a lot of skepticism, lots and lots of skepticism about things like vaccine development and getting a vaccine. And I think that was just because of differences in education or possibly community dynamics that I’m not privy to, but definitely those affected my ability to provide inpatient care.*” (6)

> “*I actually had a patient who happened to be African American who was actually in medicine, who was pregnant, and she was just like, don’t talk to me about it again. I feel like I’m being profiled and I’m not getting the vaccine. I’m not gonna do it. She’s like, nobody knows enough about this, everybody thinks they do, but I feel like, she’s like, I feel like I’m being racially profiled, because you’re pointing it out. I’m like, actually, well, I’m asking everybody about their vaccination status, but that, the tenor of it was not something that that had ever been really directed at me by a patient like that I would be racially profiling them for not. So. Yeah.*” (5)

Additionally, discussions and policies related to the pandemic, such as mask-wearing and telehealth, generated frustrations among patients, leading to their refusal to engage with MCPs regarding vaccination. MCPs reported that some patients expressed anger and indifference towards COVID-19 vaccination, stating that they did not want to hear about it or be pressured to receive it. One MCP recounted patients being frustrated or bemused by the various policies implemented, while another shared an experience with a pregnant AA patient who felt racially profiled and refused vaccination.

> “… *for my other patients, um where I tended to see it was, they were either frustrated or bemused either by all of these policies and things that were being put into place like masking and telehealth and waiting in your car after you check in that sort of thing. Or they were angry, that we kept talking about things like vaccine in particular is a big example. I got lots of people who were like, stop talking to me about this. ‘I don’t care about this.’ ‘I don’t want to hear about it.’ ‘I don’t care’.*” (6)

> “*And then, you know, I had, I actually had a patient who happened to be African American who was actually in medicine, who was pregnant, and she was just like, don’t talk to me about it again. I feel like I’m being profiled and I’m not getting the vaccine. I’m not gonna do it. She’s like, nobody knows enough about this, everybody thinks they do, but I feel like, she’s like, I feel like I’m being racially profiled, because you’re pointing it out. I’m like, actually, well I’m asking everybody about their vaccination status, you know, but that, the tenor of it was not something that that had ever been really directed at me by a patient like that I would be racially profiling them for not. So. Yeah.*” (5)

### 3.4 Concerns regarding pre-existing health conditions and potential side effects of the vaccines

Some AA women expressed concerns regarding pre-existing health conditions and potential side effects of the COVID-19 vaccines, which contributed to their hesitancy and stress when deciding whether to get vaccinated. These pregnant women expressed a desire to mitigate any potential risks that the vaccine might pose to their body and pregnancy. One woman highlighted the stress of considering vaccination after having already contracted COVID-19 multiple times. Another woman mentioned a history of Guillain-Barré syndrome, which raised concerns about the potential adverse effects of vaccination. In addition, a woman shared worries about her past experiences with vaccines, experiencing severe illness after receiving the flu shot, and having asthma.

> “*It was stressful just trying to think about whether or not I wanted to do that [get vaccinated], did I want to go through that [vaccination]? Because I did have it [COVID] already, twice.*” (1)

> “*I can’t have any vaccines. I have a history of the Guillain-Barré syndrome… But because I haven’t had a vaccine since I was an elementary school and I didn’t know if I took that right then and there if it would land me in the hospital. I said I just couldn’t do it. I couldn’t take that risk.*” (3)

### 3.5 Emotional factors associated with vaccination decision-making processes

> AA women expressed fear or concerns regarding the COVID-19 vaccine, and MCPs echoed these sentiments in their recollection of patient observations. The main fears and concerns came from worries about potential side effects, introducing something new to the body, and the long-term effects on themselves and their infants. The decision to weigh the risk of contracting COVID-19 against the vaccine proved to be challenging for many women.

> “*My husband recently got his first dose maybe three or four weeks ago, and he felt bad. I just don’t want to feel that. So I’m kind of nervous about the side effects, honestly. I am going to get my vaccine shot, but I’m just kind of mentally preparing to do that because I don’t know what could happen.*” (1)

> “*It can be really scary just to introduce anything new to your body, and making that decision to be vaccinated, I think was a really big one for a lot of moms. But I think that was difficult, I think, weighing the risk of getting COVID versus getting the vaccine and having to decide just weigh that risk.*” (9)

> “*I get a lot of patients who are nervous about the covid vaccine in pregnancy, even though we try to provide them with as much data as we can. Because it’s a relatively new vaccine. And because we know the health care benefits to moms who don’t get sick with COVID. Patients are still really scared about taking a vaccine that has not been studied for years and years. And they’re worried about the long-term effects on the baby, long term effects on them, if they go ahead and get this vaccine. But to me, it’s like, well, I don’t understand that in respect to why do you want to get the illness instead? And then they’re like, well, that’s more natural.*” (1)

Some AA women expressed ambivalence about vaccination, neither strongly for nor against it, with concerns or uncertainties about vaccines. They described a sense of wariness and a lack of anxiousness or opposition toward getting vaccinated.

> “*I don’t want to say anxiety or concern. I think that I take a stand of being weary of it, but not against it. I thought maybe there was a possibility that they would say, before we left the hospital, we’re gonna vaccinate her. I didn’t know how that was gonna go. Or if they were going to require it going into the hospital, I wouldn’t have been against it. Either way, just because I wasn’t that person that was against the vaccine. But it didn’t add any extra anxiousness or anything like that.*” (0)

> “*I was trying to figure out, did I need to get it? Or should I get it? But I didn’t get it.*” (7)

For some other AA women, the decision to receive the COVID-19 vaccine was less about choice and more about mandates imposed by their medical or work situations. Although they may have initially felt skeptical or hesitant, the requirement compelled them to proceed with vaccination. A few participants expressed their internal conflict.

> “*Honestly, it bothered me a lot. Because when they first started, doing the vaccines, everybody had all these negative things to say about it, and how they felt like, it wouldn’t work. … It’s like I always been kinda skeptical. I was like, Lord, if I don’t have to get it, then I’m fine. But because I was in hospital, I think two and a half weeks before I had him. I had no choice but to get it.*” (2)

> “*I already had in my mind that I really didn’t want to get the vaccine. But because it was just kind of like so many different emotions and feelings about the vaccine itself, drew me to my decision. … and my employer required us so that that’s the only reason why I did it.*” (4)

### 3.6 Concerns about the well-being of their infants

Some participants expressed the motivation to vaccinate their children, primarily driven by concerns for their safety and well-being. One woman highlighted the importance of vaccination due to her child attending school, where mask policies were not strictly enforced. One AA woman mentioned vaccinating their children while acknowledging their own decision not to receive vaccines. Their motivation stemmed from wanting to prioritize their children’s safety and take proactive measures to keep them protected.

> “*The older child was vaccinated, but there was some concern because he went to school, and they weren’t as strict with the mask policy.*” (8)

> “*I let my kids get shot because they were going to school but I never got shot … so I wanted to be on the safe side when it comes to them.*” (7)

Some women expressed a lack of trust in the government and doubted the thoroughness of the research conducted for vaccines. They raised concerns about vaccines’ effectiveness and safety, despite acknowledging the risks associated with contracting COVID-19. This skepticism led them to weigh the pros and cons, ultimately deciding against vaccinating their children.

> “*Because I really don’t trust. And I know I sound crazy. I don’t trust the government … [I know] I had a baby on Medicaid, but I don’t trust the government for that vaccine, I don’t feel like there was too much research done. It was, you know, they came up with it so fast. So I didn’t know I didn’t trust the um effectiveness of it. And the safety-ness of it, even though I mean, catching COVID, vaccine. So I kind of weighed out the pros and cons as far as my current health status, and the likelihood of me um of COVID being fatal if I did catch it. So I kind of just um did not get the vaccine for me or my children.*” (1)

### 3.7 Cultural perspectives

Several participants stated religious beliefs or a lack of belief in the use of vaccines. MCPs mentioned encountering postpartum patients who firmly stated that they did not believe in vaccination, attributing it to their religious beliefs. They expressed a reluctance to participate in what they perceived as an experimental practice. One MCP highlighted that although there was no official backing from religious authorities such as the Pope or prominent Islamic scholars, women held strong convictions that prevented them from considering new information and changing their stance on vaccination.

> “*I remember rounding on them [COVID-19 vaccines]and going down to postpartum. And the vaccines weren’t new, they’ve been out for a bit, but trying to say to my postpartum, ‘while you’re here, can I vaccinate you for COVID?’ And being told again, and again. No, I don’t do that. That’s not something I do. That’s not something I believe in. I don’t want to be an experiment.*” (6)

> “*Because people bring up like religious beliefs or something but that’s really not backed up in any by any sort of person that’s high up and like, it’s not backed up by the Pope. It’s not backed up by any other, religious scholar in the Islamic world. But people have their convictions that they’re not really willing to forego when they see new information.*” (8)

### 3.8 Encouragement by trusted supporters

Trusted supporters, including health care providers and family members, played a crucial role in encouraging women to receive vaccines. Their influence was significant in shaping the decision-making process, as participants relied on their advice and expertise. Participants recounted instances where their mothers or health care providers positively influenced their decision to get vaccinated.

> “… *my mom, she’s a CNA nurse, and she works in a lot of the elderly departments … and so she was already on top of it, getting all the information and she was like, ‘Look, go talk to your doctor. I don’t know if it’s okay for you to do it. But I would rather you and baby be okay’. Because of all of this, I went and talked to the doctor and they were like, ‘Yes, it’s okay. The antibodies will transfer to baby’. And I was like, okay, sign me up.*” (9)

> “*Again, my first appointment, my first postpartum appointment back. And she was just like, ‘[participant’s name], things are getting real. And so I know you were on the fence about getting this vaccine. But if you get this vaccine while you’re breastfeeding, the baby’ll get antibodies too. So you know, I wouldn’t tell you anything that I’m not telling anybody else. I wouldn’t tell you to get it’. And I haven’t gotten it [COVID]. And so she was like, ‘I really think you should get it because things are getting crazy’. And so I went ahead and got it [vaccination].*” (8)

## 4. Discussion

This study explored the factors that influence decision-making regarding COVID-19 vaccination among AA and Hispanic pregnant and postpartum women. Barriers influencing vaccination decision-making included awareness of health threats associated with COVID-19 vaccines, vaccine availability and accessibility, vaccine-related knowledge and exposure to misinformation, concerns about pre-existing health conditions and potential side effects of the vaccines, emotional factors associated with vaccination decision-making processes, concerns about the well-being of infants, and different cultural perspectives. Encouragement from trusted supporters acted as a facilitator for women’s vaccination decision-making.

Participants expressed awareness of the health threats associated with COVID-19 vaccines and concerns about the risks to themselves and their infants. This finding complements previous research highlighting the heightened awareness and concerns about the health threat posed by infectious diseases during pregnancy, such as the influenza virus (18). In addition, the availability and accessibility of COVID-19 vaccines emerged as crucial factor in women’s vaccination decision-making process. This finding reflects the early stages of the pandemic when vaccine availability was limited, and there was high demand for vaccines (19). Similar challenges were observed during the initial distribution of the influenza vaccine in the H1N1 pandemic, where demand initially exceeded supply (20). The accessibility of COVID-19 vaccines was influenced by multiple factors, including difficulties in accessing vaccination sites or lack of convenient options, particularly among underserved populations (21). Moreover, participants expressed a mix of anticipation, anxiety, and frustration related to the availability of vaccines and the challenges encountered in obtaining necessary supplies and appointments. Enhancing accessibility through initiatives such as mobile vaccination clinics, community outreach programs, and targeted scheduling systems can help alleviate the challenges expressed by participants and contribute to increased vaccination.

Misinformation and skepticism surrounding COVID-19 vaccines pose significant barriers to pregnant women’s vaccination decision-making, underscoring the importance of addressing vaccine-related knowledge gaps and countering exposure to misinformation. Social media platforms have been identified as sources of exposure to misinformation, conspiracy theories, and conflicting information about vaccines (11). The ease of sharing and accessing information on social media can lead to the rapid dissemination of misleading content, undermining public trust, and contributing to vaccine hesitancy.

Participants also raised concerns about potential side effects and the desire to observe long-term safety data. These concerns are not unique to the COVID-19 vaccine and have been observed on various vaccines (22). Addressing misinformation and leveraging social media platforms as channels for accurate and evidence-based vaccine information can be a promising approach to promote vaccination. Providing accurate information about vaccine safety, addressing misconceptions about side effects, and alleviating concerns about long-term safety are crucial for increasing vaccine acceptance among pregnant women.

Emotional factors, such as fear of side effects and apprehension about introducing something new to their bodies, contribute to the vaccination decision-making process among pregnant women. The unique physical and emotional demands of pregnancy and the postpartum period intensify these factors (23–26). The decision-making process during pregnancy and postpartum period is accompanied by heightened responsibilities, as women consider the potential impacts on both their own health and the well-being of their babies. The weight of these responsibilities may exacerbate the emotional concerns and anxiety associated with vaccination decision-making, as women strive to make choices that prioritize the safety and health of both themselves and their babies.

Cultural perspectives emerged as significant factors affecting the COVID-19 vaccination decision-making process among AA and Hispanic women. Previous studies have highlighted the importance of implementing culturally tailored communication strategies and multilingual resources to improve vaccine acceptance in diverse populations (27). It is worth noting that all Hispanic participants in our study were immigrants, and 70% relied on emergency Medicaid to cover childbirth expenses. These findings highlight the need to address the specific challenges faced by immigrant populations and the importance of providing appropriate support in accessing health care services during the pandemic.

Moreover, the level of trust in the health care system and government emerged as a crucial determinant of vaccine acceptance. Building trust through transparent communication, community engagement, and addressing historical concerns is vital for improving vaccine confidence among AA and Hispanic women. Trusted supporters, such as health care providers and family members, play a crucial role in encouraging pregnant and postpartum women to get vaccinated against COVID-19. Trusted supporters can provide essential information, address concerns, and offer emotional support, thereby influencing decision-making processes.

MCPs, in particular, hold a position of trust and authority in maternal health care, making them key influencers in promoting vaccination among pregnant and postpartum women. Engaging in a conversation with MCPs may assist in decisions regarding COVID-19 vaccination during pregnancy. Conversations about risk should take into account the individual patient’s values and perceived risk of various outcomes (28). Empowering MCPs through education on the importance of COVID-19 vaccination and providing them with accurate information can enable them to effectively communicate the benefits and safety of vaccines to their patients. Moreover, involving family members and community leaders in health communication campaigns can leverage their influence to promote vaccination and address any misconceptions or fears (11). By harnessing the power of trusted supporters, health care systems can enhance vaccine acceptance among AA and Hispanic pregnant and postpartum women, ultimately contributing to better health outcomes for both mothers and their babies.

There are several limitations to this study. First, this study was based on a broader project. The interview guide used for the broader project did not focus on specific vaccination-related topics, which may have influenced the depth of participants’ responses. Second, the Hispanic participants in this study were all immigrants, which may limit the generalization of the finds to Hispanic mothers with different background. Third, we did not recruit white women in this study; thus, we cannot direct compare the experiences of different racial and ethnic groups in vaccination decision-making. Future studies should consider diverse samples to further explore these factors and develop targeted interventions to improve vaccine acceptance in these populations.

## 5. Conclusion

COVID-19 vaccination reduces the risk of severe illness and death from COVID-19, but AA and Hispanic pregnant and postpartum women face unique challenges that affect their vaccination decision-making. Women’s lived experiences and MCPs’ observations reveal barriers such as fear of potential vaccine side effects, misinformation, and distrust of the health care system prevented these women from vaccination. Conversely, reliable information, social support, and trusted doctors’ advice can be facilitators for promoting COVID-19 vaccination. Future interventions should prioritize addressing health care disparities among pregnant women of color by actively involving trusted doctors in their communities, disseminating accurate information, and initiating tailored vaccination conversations during pregnancy and postpartum care. Moreover, providing insurance support for Hispanic women with immigrant backgrounds can address the challenges they faced during the pandemic. By recognizing and addressing these challenges, public health professionals can develop targeted communication strategies and interventions to improve vaccine acceptance and contribute to better health outcomes for AA and Hispanic pregnant and postpartum women.

## Author Contributions

JL and SQ conceptualized and designed the study. RZ and TB wrote the first draft and SQ, JL, MET, and XL participated in writing sections of the original proposal. All authors critically reviewed and edited the manuscript. JL, MET, TB, and RZ participated in data collection, data analysis, and data interpretation. JL and XL secured the funding.

## Institutional Review Board Statement

The study was approved by the Institutional Review Board of the University of South Carolina (Pro00115169).

## Acknowledgements

We express our gratitude to the participants for their willingness to share their lived experiences and for dedicating their time to the interviews.

## Informed Consent Statement

Informed consent was obtained from all subjects involved in the study.

## Data Availability Statement

The data presented in this study are available on request from the corresponding author. The data are not publicly available due to privacy concerns.

